# Dissecting shared genetic architecture between hypothyroidism and immune-related diseases

**DOI:** 10.1101/2024.04.02.24305135

**Authors:** Shifang Li, Meijiao Gong

## Abstract

Hypothyroidism is a common endocrine disorder characterized by insufficient thyroid hormone production and there are increasing reports of hypothyroidism being associated with a variety of complex diseases. However, whether these associations share a common genetic basis remains unclear. In this study, we investigated the shared genetic architecture underlying hypothyroidism and three immune-related diseases, including sarcoidosis, chronic sinusitis, and interstitial lung disease (ILD) endpoints, using large-scale genome-wide cross-traits analysis. Linkage disequilibrium score regression and genetic covariance analysis revealed significant genetic correlations between hypothyroidism and each of the three diseases. Cross-trait meta-analyses identified 26 shared risk loci between hypothyroidism and other three diseases. Cell-type SNP heritability enrichment analyses highlighted that central memory CD4⁺ T cells in the blood are co-enriched by these four diseases, which was further confirmed by co-localization analysis (posterior probability>0.9). Furthermore, DOCK6 and CD226 were identified as candidate causal genes shared by hypothyroidism and sarcoidosis, while RIPK2 was found as a hypothyroidism-driven plasma protein that may mediate the risk of ILD endpoints. Overall, these findings provide new insights into the shared genetic underpinnings of hypothyroidism and immune-related diseases and offer potential targets for comorbidity risk assessment and therapeutic intervention.

## Introduction

Hypothyroidism is an endocrine disorder characterized by deficient synthesis, secretion, or action of thyroid-stimulating hormone (TSH), resulting in impaired thyroid function^1^. The regulation of TSH is known to be polygenic in nature, with recent genome-wide association studies (GWAS) identifying 42 loci that collectively account for approximately 33% of the genetic variance in TSH levels^2^. While intrinsic factors such as age and sex, as well as environmental influences including smoking and body mass index (BMI), also modulate thyroid function, their combined explanatory power remains limited, accounting for only 7% of the variance in TSH levels and 5% in free thyroxine concentrations^3,4^. Notably, the burden of hypothyroidism is disproportionately higher among women, with prevalence rates approaching 7% in those aged 85 to 89 years^5–7^.

Beyond its primary clinical manifestations, hypothyroidism has been increasingly linked to a range of complex diseases. For instance, epidemiological evidence suggests an elevated risk of dementia among older adults with hypothyroidism^7^, and clinical observations have noted associations with sarcoidosis and chronic rhinitis^8,9^. Furthermore, in interstitial lung diseases (ILD), particularly idiopathic pulmonary fibrosis and fibrosing hypersensitivity pneumonitis, hypothyroidism has been associated with poorer survival outcomes^10,11^. However, whether these associations reflect causal relationships or arise due to residual confounding remains unclear, as observational studies are inherently vulnerable to bias from unmeasured variables.

To address this challenge, Mendelian randomization (MR) offers a genetically informed approach to assess causal relationships by using genetic variants as instrumental variables^12^. Despite the increasing availability of GWAS data for hypothyroidism and various complex traits, the extent to which these conditions share a genetic basis is not well characterized. Cross-trait meta-analysis approaches have been widely employed to uncover shared genetic variants among diseases such as obesity and multiple sclerosis^13^, gastrointestinal disorders and endometriosis^14^, and autoimmune diseases including multiple sclerosis and inflammatory bowel disease^15^, underscoring the value of investigating pleiotropic genetic effects.

In this study, we utilized publicly available GWAS summary statistics derived from individuals of European ancestry to investigate the genetic correlation and causality between hypothyroidism and three immune-related diseases: sarcoidosis, chronic sinusitis, and interstitial lung disease (ILD) endpoints (**Figure 1**). In addition, a large-scale, genome-wide cross-trait analysis was conducted to investigate the potential shared genetic components in these diseases. The biological consequences revealed by common loci could play key roles in the co-occurrence of hypothyroidism and immune-related diseases.

**Figure 1.**
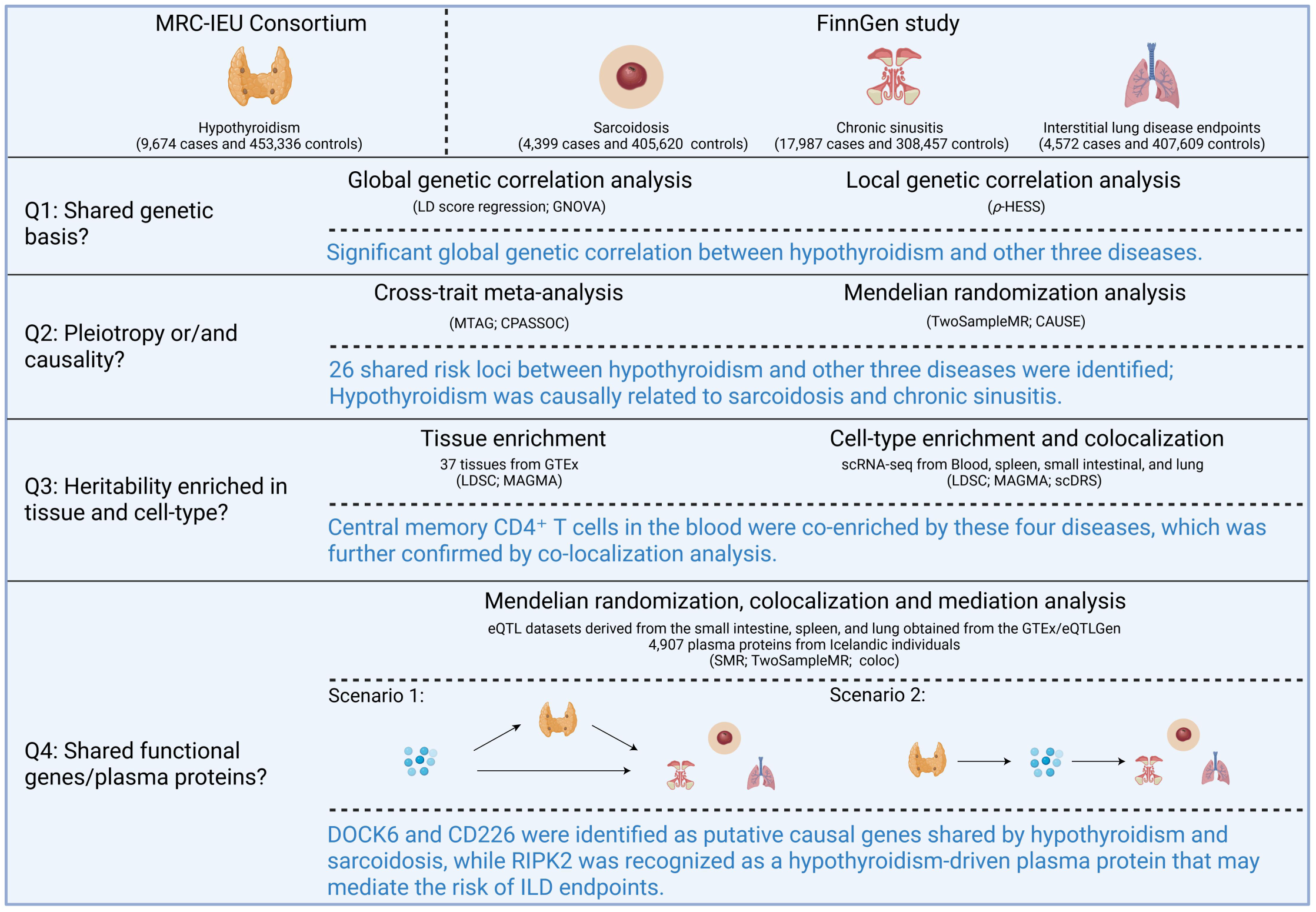
Study overview and summary. CPASSOC: Cross Phenotype Association; GNOVA: Genetic covariance analyser; LDSC: linkage disequilibrium score regression; MAGMA: Multi-marker Analysis of GenoMic Annotation; scDRS: single-cell Disease Relevance Score; MTAG: Multi-Trait Analysis of GWAS; MR: Mendelian Randomization; ρ-HESS: Heritability Estimation from Summary Statistics; CAUSE: Causal Analysis Using Summary Effect; scRNA-seq: single-cell RNA sequencing; SMR: Summary-data based Mendelian randomization.

## Results

### Significant genetic correlation between hypothyroidism and immune-related diseases

Bivariate linkage disequilibrium Score Regression (LDSC) was first employed to assess the genome-wide genetic correlation between hypothyroidism and three immune-related diseases (sarcoidosis, chronic sinusitis, and ILD endpoints), without constraining the regression intercept (**Figure 2A**). This unconstrained analysis identified significant genetic correlations between hypothyroidism and ILD endpoints (genetic correlation Rg=0.2425, *p=*0.0063), sarcoidosis (Rg=0.2937, *p*=2.05×10^−5^), and chronic sinusitis (Rg=0.1776, *p*=0.0004). Liability-scale SNP heritability estimates were calculated as 0.0469 for hypothyroidism, 0.0042 for ILD endpoints, 0.0126 for sarcoidosis, and 0.0241 for chronic sinusitis. The LDSC intercepts, ranging from 0.018 to 0.040, indicated minor sample overlap between hypothyroidism and other three diseases. Given the minimal sample overlap between the MRC-IEU Consortium^16^ and FinnGen dataset^17^, the intercepts were subsequently set as zero to enhance statistical power and reduce standard error in the LDSC estimates. This adjustment yielded slightly attenuated, though still statistically significant, genetic correlations (**Figure 2A**). Additional analyses using GeNetic cOVariance Analyzer (GNOVA)^18^ and Heritability Estimation from Summary Statistics (ρ-HESS)^19^ further corroborated the presence of robust genetic associations between hypothyroidism and each of the three immune-related diseases (**Figure 2A**).

**Figure 2.**
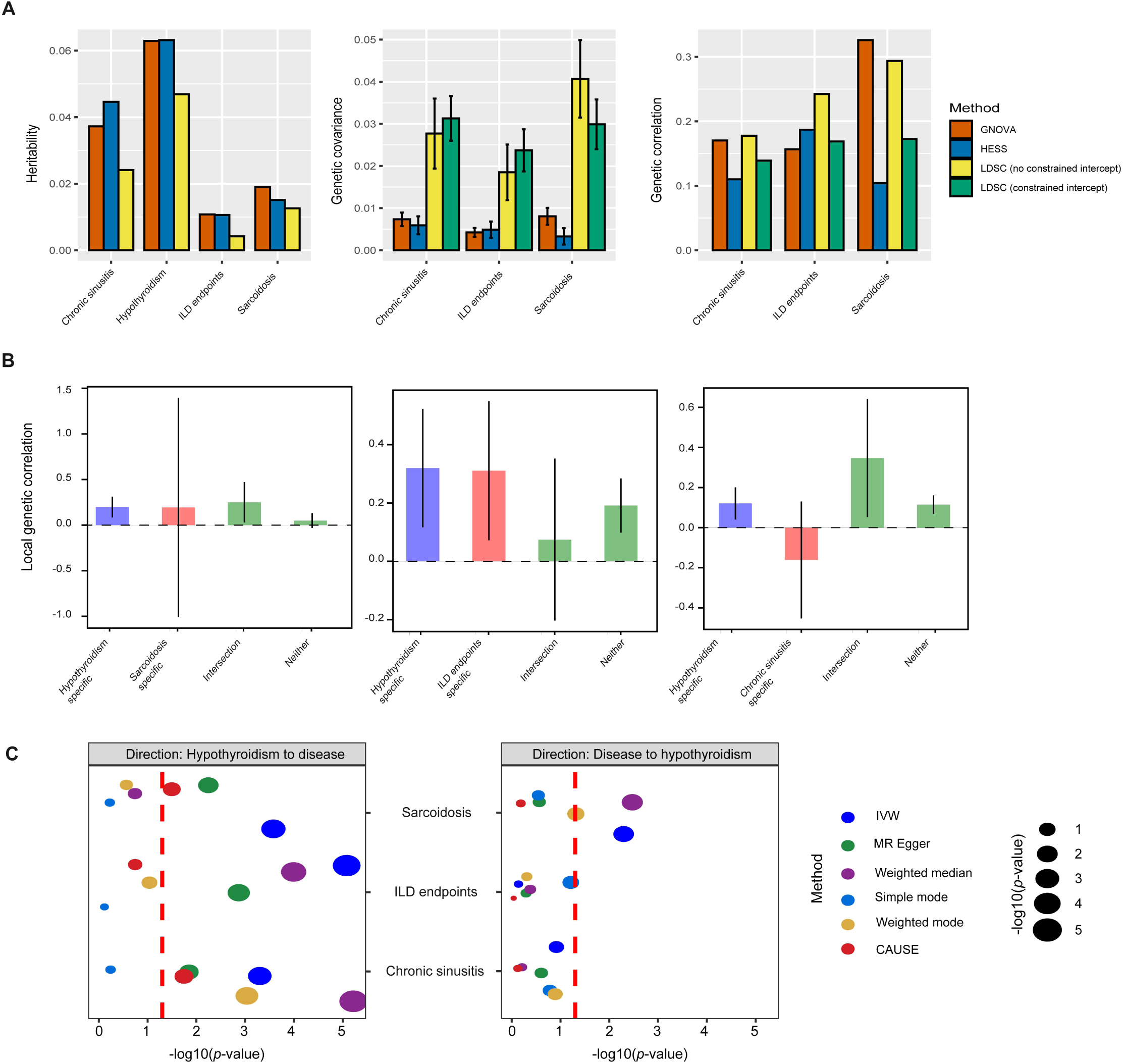
Genetic correlation and causality between hypothyroidism and three immune-related diseases. (**A**) The heritability, genetic covariance, and global genetic association of hypothyroidism with three immune-related diseases. (**B**) The average local genetic association between hypothyroidism and three immune-related diseases. The error bars show the 95% confidence intervals of the estimates. (**C**) Summary of bidirectional MR analyses of hypothyroidism and three immune-related diseases. The abbreviation IVW refers to inverse variance weighting. The dot represents -log10 *p*-values in MR analysis. The red dotted line represents the -log10(0.05). LDSC, linkage disequilibrium score regression; GNOVA, genetic covariance analyzer.

To explore whether these associations were attributable to specific genomic loci, local genetic correlation was assessed at GWAS loci relevant to each disease. Thirteen region pairs showed suggestive significance (uncorrected *p*<0.05, ρ-HESS; **Supplementary Table S2**), comprising four regions shared between hypothyroidism and chronic sinusitis, six between hypothyroidism and sarcoidosis, and three between hypothyroidism and ILD endpoints. Mean local genetic correlations were comparable across loci specific to hypothyroidism and those specific to the other diseases (**Supplementary Figure S1**). No additional regions exhibited substantial local genetic covariance. Notably, loci associated with hypothyroidism showed stronger local genetic correlations than those unique to sarcoidosis (local Rg=0.2, standard error [SE]=0.058 *vs* local Rg=0.19, SE=0.61) or chronic sinusitis (local Rg=0.12, SE=0.041 *vs* local Rg=-0.16, SE=0.15) (**Figure 2B**), suggesting that alleles conferring increased susceptibility to hypothyroidism are also likely to elevate the risk for sarcoidosis and chronic sinusitis.

Collectively, these findings imply that the observed comorbidity is largely attributable to genome-wide polygenic overlap rather than confined to discrete genomic intervals.

### Hypothyroidism is causally related to sarcoidosis and chronic sinusitis

Bidirectional MR analyses were then conducted to assess potential causal relationships and to determine whether the shared genetic architecture between hypothyroidism and the selected complex diseases could be attributed to pleiotropy. Hypothyroidism was significantly associated with the elevated risk for all three diseases: ILD endpoints (inverse-variance weighted [IVW] β=3.49, SE=0.68, *p*=2.63×10^−7^), sarcoidosis (IVW β=4.26, SE=0.87, *p*=8.83×10^−7^), and chronic sinusitis (IVW β=1.78, SE=0.47, *p*=1.87×10^−4^) (**Supplementary Table S3**). The MR-Egger intercept test indicated no evidence of directional pleiotropy, thereby supporting the robustness of the causal inference. Furthermore, leave-one-out sensitivity analysis confirmed that the observed associations were not disproportionately influenced by any single instrumental variant.

In the reverse MR analysis, no significant causal effect was detected from any of the three immune-related diseases on hypothyroidism. Specifically, the odds ratios (ORs) were as follows: ILD endpoints, OR=1.0026 (95% confidence interval [CI]: 0.998-1.0064, *p*=0.19); sarcoidosis, OR=1.0062 (95% CI: 0.99-1.012, *p*=0.061); and chronic sinusitis, OR=1.012 (95% CI: 0.99-1.03, *p*=0.209). These null findings were consistent across multiple MR methodologies (**Supplementary Table S3**).

To further substantiate these results, replication analyses were performed using the largest GWASs of hypothyroidism to date, encompassing 51,194 cases and 443,383 controls from Finnish and UK Biobank cohorts (**Figure 2C** and **Supplementary Table S4**)^20^. All previously identified causal relationships were reproduced in this independent dataset. However, the Causal Analysis Using Summary Effect estimates (CAUSE) model^21^ indicated that, in addition to ILD endpoints, only chronic sinusitis and sarcoidosis showed genetic susceptibility mediated by hypothyroidism.

### Cross-trait meta-analysis and pleiotropic loci

In light of the substantial genetic correlations observed between hypothyroidism and the three immune-related diseases, a cross-trait meta-analysis was conducted to detect the shared genetic variants. This analysis identified 26 single nucleotide polymorphisms (SNPs) reaching genome-wide significance (*p*<5×10^−8^) using both the Multi-Trait Analysis of GWAS (MTAG)^22^ and Cross-Phenotype Association (CPASSOC) methodologies^23^ (**Figure 3**; **Supplementary Table S5**). The maximum false discovery rates (maxFDR) from the MTAG analyses were 0.00044 for hypothyroidism, 0.00825 for sarcoidosis, 0.026 for chronic sinusitis, and 0.27 for ILD endpoints. Furthermore, the high concordance between results from MTAG and CPASSOC supports the reliability of the findings and suggests minimal bias arising from MTAG model assumptions.

**Figure 3.**
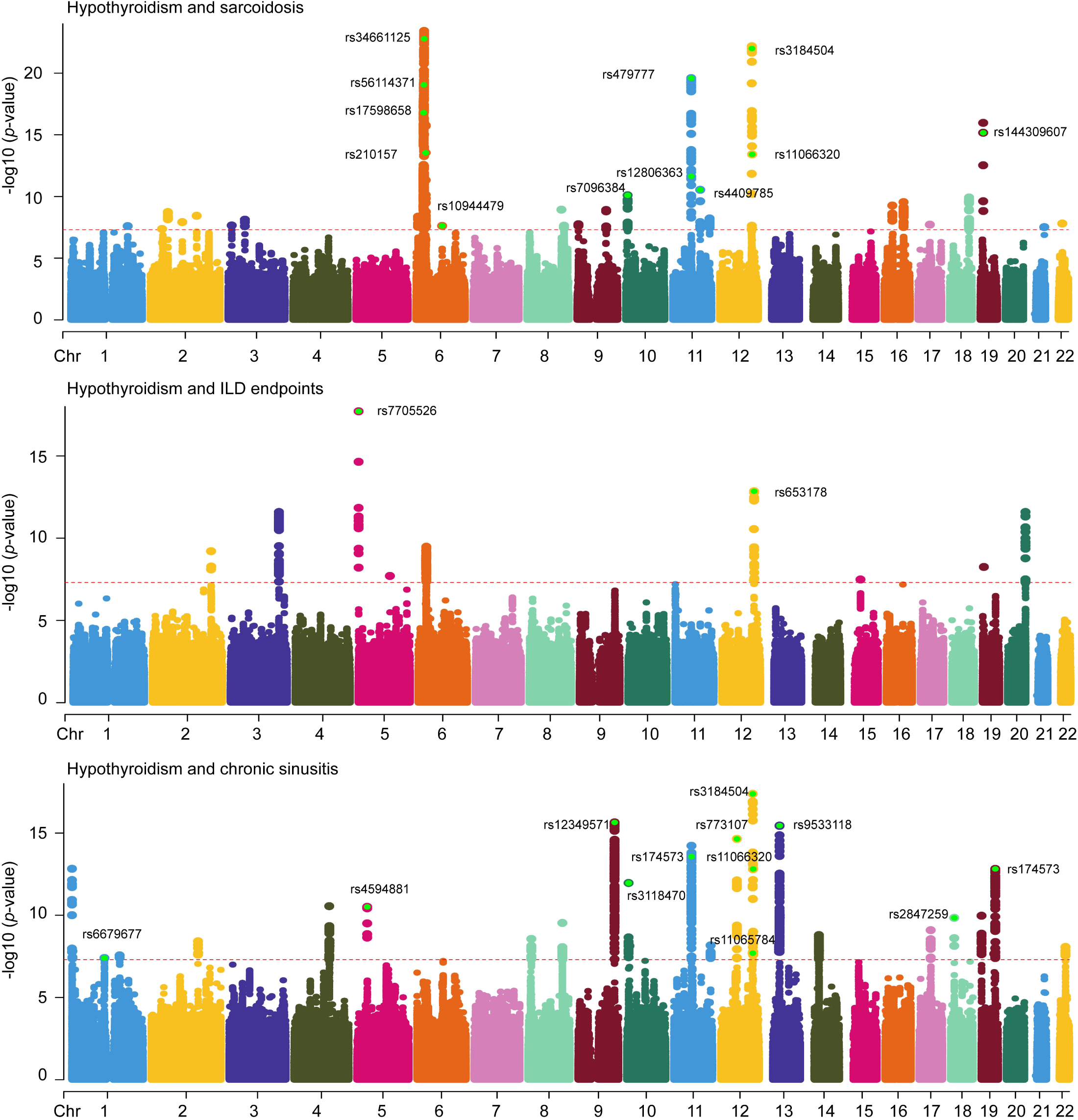
Manhattan plot for genome-wide association study using cross-trait GWAS meta-analysis. The x-axis displays the number of autosomes, while the y-axis displays the -log10 *p*-values for statistical significance derived from the CPASSOC model. The dots represent SNPs. The red dotted line represents the genome-wide significance threshold (*p*<5e-08). Pleiotropic loci that are both significant in single-trait and cross-traits GWASs, including MTAG and CPASSOC model, were indicated by text.

Of the 26 significant loci, 12 loci were associated with both hypothyroidism and chronic sinusitis, two with hypothyroidism and ILD endpoints, and 12 with hypothyroidism and sarcoidosis. Notably, two novel associations were identified: one between hypothyroidism and chronic sinusitis at rs174573 (closest gene FADS2, *p-*_hypothyroidism_=1.30×10^−7^, *p-*_chronic sinusitis_=1.04×10^−7^, *p-*_CPASSOC&MTAG_<5×10^−8^), and the other between hypothyroidism and sarcoidosis at rs12806363 (closest gene RPS6KA4, *p-*_hypothyroidism_=6.4×10^−8^, *p-*_sarcoidosis_=1.52×10^−6^, *p-*_CPASSOC&MTAG_<5×10^−8^). Of note, 11 of these 26 SNPs remained statistically significant (*p*<0.05) under local genetic correlation analysis using the ρ-HESS approach (**Supplementary Table S2**) while 16 of these 26 SNPs exhibited strong colocalization probabilities (PH4>0.8), indicating that the same causal variants likely underlie the associations with both hypothyroidism and the other diseases (**Supplementary Table S5**). Several of these loci were consistently validated across multiple analytical approaches. Specifically, two SNPs (rs11066320 and rs3184504) shared between hypothyroidism and sarcoidosis; one SNP (rs653178) shared with ILD endpoints; and four SNPs (rs12349571, rs11065784, rs11066320, and rs3184504) shared with chronic sinusitis showed colocalization probabilities above 0.8 (**Supplementary Table S6**). Interestingly, the loci rs11066320 and rs3184504, which were jointly associated with hypothyroidism and sarcoidosis, also emerged in the analysis of hypothyroidism and chronic sinusitis (**Supplementary Table S5**). Furthermore, rs653178, shared between hypothyroidism and ILD endpoints, is located within an intronic region of ATXN2, a site that also functions as a transcription factor-binding region for XBP1, CREB3, BATF3, JDP2, FOS, ATF7, and JUN.

To prioritize likely causal variants at each pleiotropic locus, credible variant sets were generated based on posterior probabilities^24^, retaining SNPs with a cumulative probability of 95% (**Supplementary Table S7**). In total, 147 candidate variants were identified. Among these, five SNPs (rs11066320, rs2847259, rs3184504, rs6679677, and rs7705526) exhibited posterior probabilities exceeding 0.99, suggesting high confidence in their potential causal roles. These credible sets provide valuable targets for future functional validation studies.

### SNP heritability enrichment at the tissue and cell-type levels

To elucidate the tissue-specific genetic architecture underlying hypothyroidism and related complex diseases, we employed LDSC and Multi-marker Analysis of Genomic Annotation (MAGMA)^25^ to identify 37 tissues with enriched genetic associations, utilizing data from the GTEx v8 resources^26^. Both methods assessed whether the top 10% of tissue-specific genes exhibited elevated levels of genetic association with each trait. After controlling for baseline annotations and correcting for multiple comparisons, significant enrichment of SNP-based heritability for hypothyroidism was observed in four tissues-spleen, small intestine, peripheral blood, and lung (**Figure 4A**; **Supplementary Table S8**). No significant enrichment was detected for sarcoidosis, chronic sinusitis, or ILD endpoints following adjustment for multiple testing.

**Figure 4.**
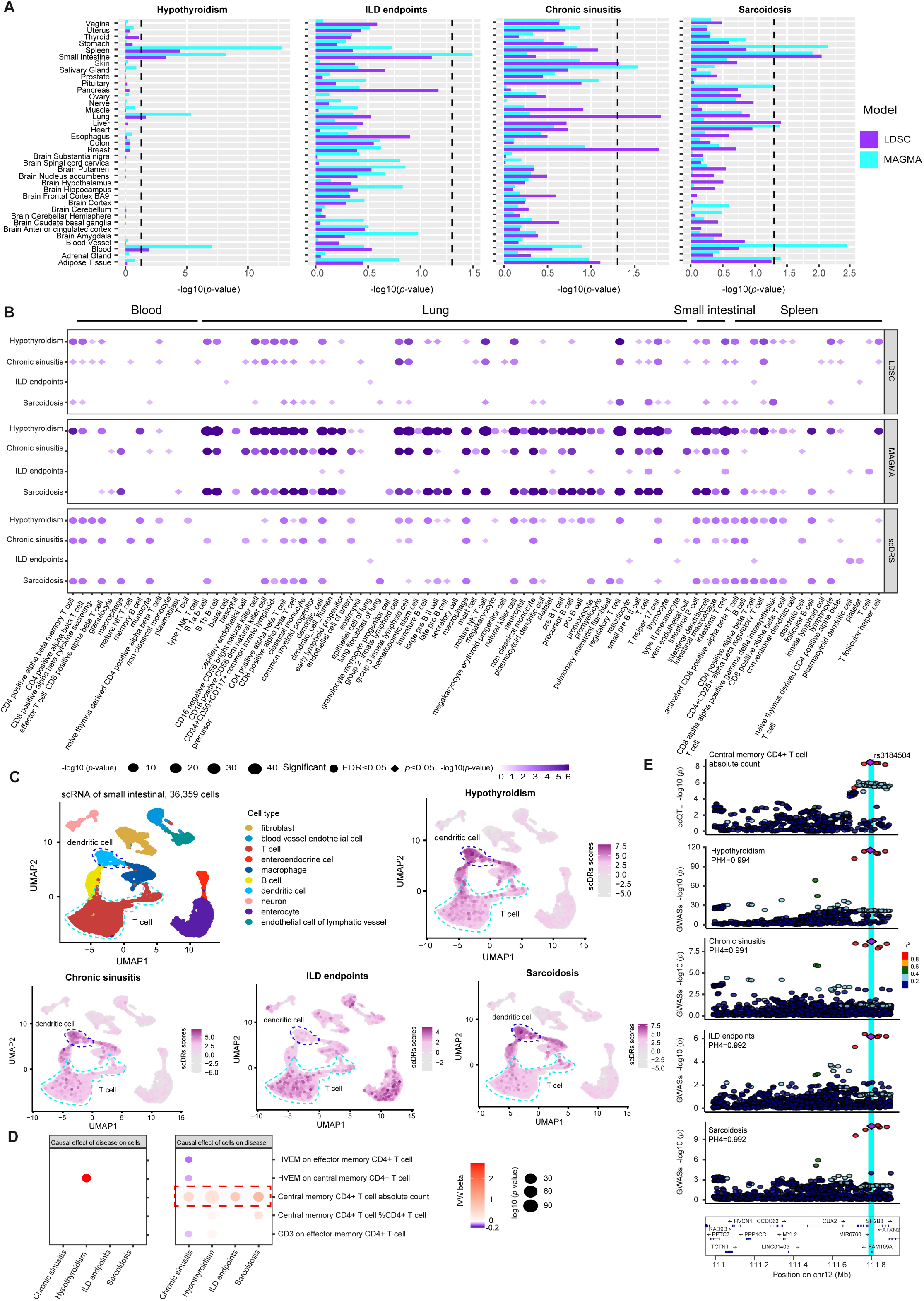
Tissue/cell type-specific enrichment of SNP heritability for four diseases. (**A**) The heritability enrichment of tissues for diseases using LDSC and MAGMA. The x-axis displays -log10 *p*-values for each individual test. (**B**) The heritability enrichment of cell types for four diseases using LDSC, MAGMA, and scDRS. An expression specificity matrix created with CELLEX was used to perform LDSC-SEG and MAGMA regression analyses. For scDRS analysis, a co-variable matrix containing assay, gender, age, ethnicity, and time of cold storage (only for the spleen scRNA dataset) was used. Dot colors and point size indicate considerable cell type-disease associations. scDRS: single-cell disease relevance score. FDR indicates that the *p*-value was adjusted using the Benjamini-Hochberg method for all cell types across diseases. (**C**) UMAP embeddings of cell types in the small intestinal with normalized scDRS scores for the four diseases. Pink represents cells that are enriched for the aforementioned disease, while gray implies non-relevant cells. (**D**) The causal association between the listed cell types and four diseases. The dot size shows -log10 *p*-values from MR analysis using the IVW approach. (**E**) Colocalization analyses of cell type QTL for central memory CD4+ T cell absolute count in four diseases. We utilized colocalization to investigate whether the QTL for central memory CD4+T cell absolute count shared the same causative variant as four other diseases. Each dot indicates a single nucleotide polymorphism, while colors indicate linkage disequilibrium (LD; r^2^) with the most likely causative variant, rs3184504. The posterior probability (PH4) was calculated using the “colco.abf” function from the coloc R package. FDR, false discovery rate.

To account for tissue heterogeneity, single-cell RNA sequencing (scRNA-seq) datasets were then analyzed from four relevant tissues, including whole blood (50,115 cells), spleen (94,256 cells), small intestine (36,359 cells), and lung (71,752 cells), to evaluate cell type-specific expression patterns associated with genetic risk for hypothyroidism and the three immune-related diseases (**Figure 4B**; **Supplementary Table S9**). Three complementary analytical approaches, including LDSC, MAGMA, and single-cell Disease Relevance Score (scDRS)^27^, were applied to robustly identify relevant cellular contexts. Particularly, in contrast to LDSC and MAGMA, scDRS enables us to detect GWAS signal enrichment at the single-cell level and quantifies intra-cell-type heterogeneity in trait associations. These analyses revealed significant enrichment of polygenic risk for hypothyroidism, sarcoidosis, and chronic sinusitis in CD4⁺ αβ memory T cells, CD4⁺ αβ T cells, and CD8⁺ αβ T cells in peripheral blood, implicating adaptive immune cell types in the shared genetic etiology of these conditions (**Figure 4B**; **Supplementary Table S9**). In the lung, hypothyroidism showed overlapping enrichment with sarcoidosis and chronic sinusitis in group 2 and group 3 innate lymphoid cells, CD4⁺ and CD8⁺ αβ T cells, regulatory T cells, and T helper 17 cells. In the spleen, activated CD8⁺ αβ T cells showed significant heritability enrichment for hypothyroidism in conjunction with sarcoidosis and chronic sinusitis. Additionally, both hypothyroidism and sarcoidosis exhibited substantial enrichment in T cells and dendritic cells within the small intestine (**Figure 4C**), suggesting shared pathogenic mechanisms at the cellular level across tissues.

Of note, heritability enrichment does not imply causality between cell types and diseases. To assess potential causal relationships, MR was also conducted using phenotypic data from 14 CD4⁺ memory T cell subtypes obtained from a Sardinian cohort of 3,757 individuals^28^ (**Supplementary Tables S10**-**S11**). After multiple testing correction, nine significant cell-disease associations were identified. In particular, the absolute count of central memory CD4⁺ T cells was genetically associated with increased risk for hypothyroidism (OR=1.008, 95% CI: 1.0029-1.013; False Discovery Rate (FDR)=1.16×10^−2^), ILD endpoints (OR=1.94, 95% CI: 1.49-2.51; FDR=9.44×10^−6^), sarcoidosis (OR=2.49, 95% CI: 1.92-3.25; FDR=1.41×10^−10^), and chronic sinusitis (OR=1.51, 95% CI: 1.32-1.73; FDR=2.86×10^−8^) (**Figure 4C**). Further colocalization analyses indicated a high probability of shared genetic variants underlying the associations between central memory CD4⁺ T cell counts and each of the four conditions: hypothyroidism (posterior probability for shared causal variant, PH4=0.994), ILD endpoints (PH4=0.992), sarcoidosis (PH4=0.992), and chronic sinusitis (PH4=0.991) (**Figure 4D**). These findings collectively suggest that hypothyroidism may play a mediating role in the observed genetic relationships between central memory CD4⁺ T cell abundance and susceptibility to immune-related diseases.

### Identification of shared functional genes for hypothyroidism and immune-related diseases

To identify candidate functional genes implicated in hypothyroidism and associated complex diseases we investigated, we integrated GWAS summary statistics with expression quantitative trait loci (eQTL) data from multiple tissues. Specifically, we utilized eQTL datasets derived from the small intestine, spleen, and lung obtained from the GTEx v8 datasets, alongside whole blood eQTL data from both GTEx and eQTLGen^29^ (**Figure 5A**; **Supplementary Table S12**). Following Bonferroni correction for multiple testing (*p*<0.05), two significant gene-disease associations, including HLA-DPB2 and HLA-DQB1-AS1, were identified in the small intestine in relation to hypothyroidism and its comorbid diseases. In the spleen, AP003774.4 and HLA-DPB2 were found to be jointly associated with both hypothyroidism and sarcoidosis. Additionally, PPP1R18, located in lung tissue, exhibited shared genetic association between hypothyroidism and sarcoidosis based on Summary-data-based Mendelian Randomization (SMR) analysis (adjusted *p-*_smr_<0.05; *p-*_Heterogeneity in Dependent Instruments (HEIDI)_>0.05), suggesting pleiotropic effects not attributable to linkage disequilibrium. Although some genes did not meet strict significance thresholds across all traits, they still showed relevance to multiple diseases. For instance, CUTALP, a gene strongly linked to hypothyroidism, was also implicated in ILD endpoints and sarcoidosis in the small intestine, whole blood, and lung. Similarly, DOCK6 exhibited associations with hypothyroidism and all three immune-related diseases in both the small intestine and blood.

**Figure 5.**
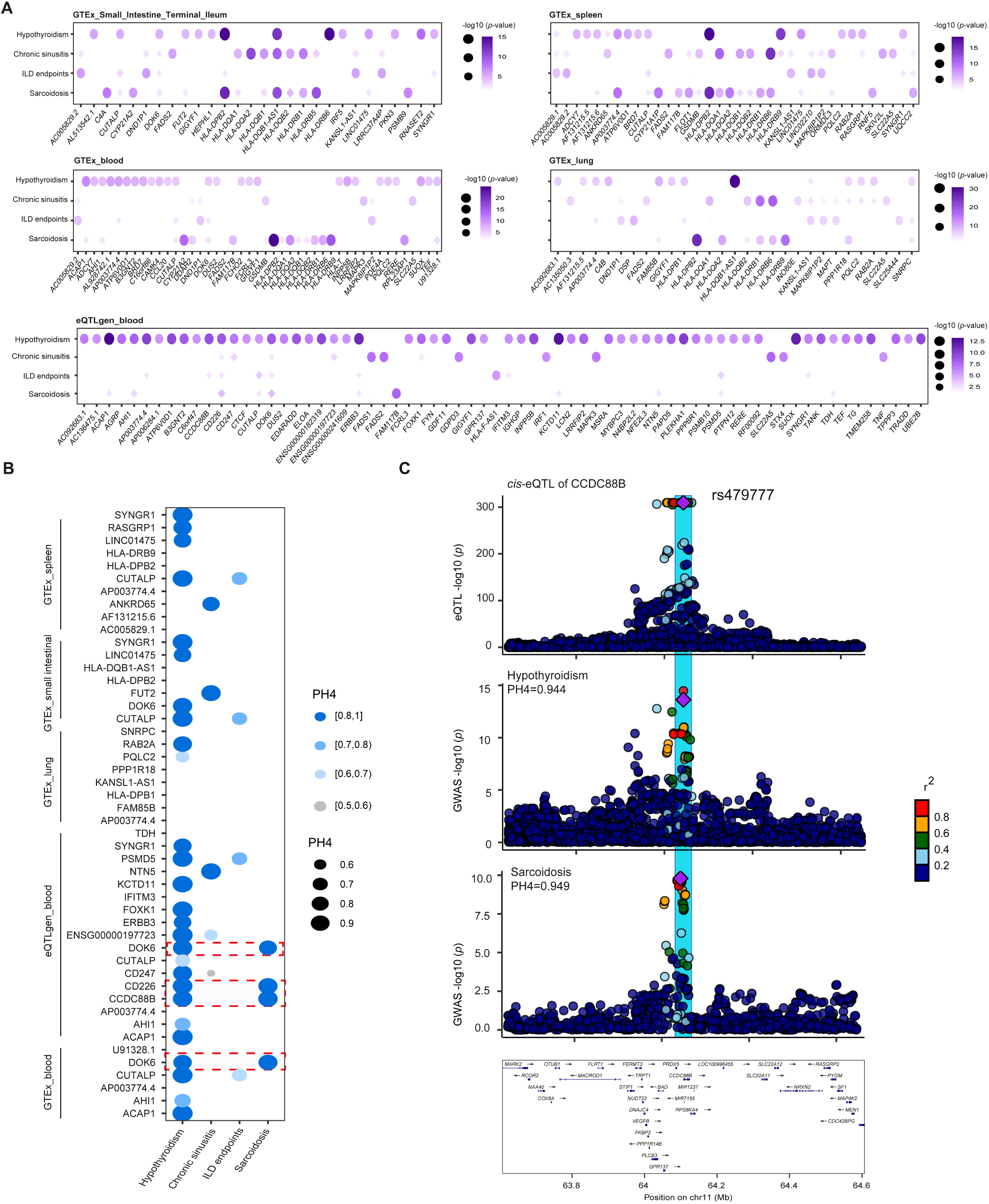
Prioritized genes related to four diseases. (**A**) SMR prioritized genes linked to four diseases. To study the association between gene expression and four diseases, *cis*-eQTL from GTEx v8 (blood, lung, spleen, and small intestine) or eQTLgene (blood) were used. Prioritized genes were statistically related to at least one disease (Bonferroni-corrected *p*<0.05), and *P*_HEIDI_ scores < 0.05 were used for plotting. The point size and color show the size of associations between genes and diseases. (**B**) Colocalization analysis of potential genes associated with four diseases. We used colocalization to determine whether the putatively shared genes identified in (A) could share the same causative variants with diseases. Colocalization analyses were performed using *cis*-eQTL from GTEx v7 (blood, lung, spleen, and small intestine) or eQTLgene (blood). (**C**) Colocalization analysis of the CCDC88B *cis*-eQTL with hypothyroidism and sarcoidosis. Each dot indicates a single nucleotide polymorphism, while colors indicate linkage disequilibrium (LD; r^2^) with the most likely causative variant, rs479777. The posterior probability (PH4) was calculated using the “colco.abf” function in coloc R package.

To further corroborate these findings, colocalization analyses were conducted across 48 gene-tissue pairs using human GTEx v7 datasets (**Figure 5B**; **Supplementary Table S13**). Notable colocalization was observed for DOCK6 and CD226 with both hypothyroidism and sarcoidosis in whole blood. Furthermore, CCDC88B showed strong colocalization with hypothyroidism and sarcoidosis in blood, sharing the genetic variant rs479777, a pleiotropic locus previously identified (**Figure 3**, **Figure 5C**, and **Supplementary Table S13**). However, while CCDC88B displayed strong evidence for association with hypothyroidism (*p-*_smr_=6.84×10^−^⁹), the association with sarcoidosis did not meet the HEIDI threshold (*p-*_HEIDI_= 6.85×10^−3^), precluding definitive inference of causality for sarcoidosis. It is also noteworthy that CUTALP, which showed strong colocalization with hypothyroidism, exhibited moderate colocalization with ILD endpoints across the small intestine, blood, and lung tissues. These results collectively suggest that shared functional genes may be associated with disease comorbidities.

### Identification of shared functional plasma proteins for hypothyroidism and immune-related diseases

Given the relevance of the plasma proteome as a rich source of potential therapeutic targets, putative circulating proteins with functional relevance shared between hypothyroidism and related complex diseases were identified using large-scale genome-wide association data on 4,907 plasma proteins derived from 35,559 Icelandic individuals^30^ (**Figure 6A**; **Supplementary Table S14**). As *cis*-acting protein quantitative trait loci (*cis*-pQTLs) are generally considered to exert more direct and specific biological effects on protein expression compared to *trans*-pQTLs, MR analyses were conducted using *cis*-pQTLs as instrumental variables and disease phenotypes as outcomes^31^. Following stringent quality control, including exclusion of instruments with evidence of heterogeneity (I^2^<50%), directional pleiotropy (MR-Egger intercept *p*>0.05), reverse causality (Steiger test *p*<0.05), and sensitivity testing via leave-one-out analysis, 34 proteins exhibited significant causal associations with disease outcomes (FDR<0.05) (**Figure 6A**; **Supplementary Table S15**). Notably, AIF1 was inversely associated with the risk of both hypothyroidism (OR, 95% CI=0.981 [0.974-0.988]; *p*=1.65×10^−7^) and chronic sinusitis (OR=0.655 [0.550-0.780]; *p*=2.00×10^−6^), despite showing a strong positive association with sarcoidosis (OR=10.33 [7.40-14.43]; *p*=6.32×10^−43^). To examine whether the effect of AIF1 on chronic sinusitis might be mediated through hypothyroidism, a two-step MR-based mediation analysis was performed using effect estimates from the initial and intermediary models^20^. The results indicated that hypothyroidism mediated approximately 7.83% of AIF1’s total effect on chronic sinusitis. Beyond AIF1, no additional proteins showed consistent statistically significant associations across both hypothyroidism and the immune-related diseases. Nevertheless, 19 protein-disease associations reached nominal significance (**Figure 6B**; **Supplementary Table S15**), of which 12 exhibited concordant effect directions between hypothyroidism and the three comorbid conditions (i.e., the product of effect estimates for protein-to-disease and protein-to-hypothyroidism was positive, and the effect estimate for hypothyroidism-to-disease was also positive). For example, genetically increased ALDH2 expression was significantly associated with higher risk of hypothyroidism (OR=1.053 [1.040-1.066]; *p*=5.26×10^−17^), as well as elevated risk for ILD endpoints, sarcoidosis, and chronic sinusitis. Additionally, elevated genetically predicted levels of ANTXR1 (OR=0.316 [0.128-0.782]; *p*=0.0127) and NBL (OR=2.704 [1.149-6.360]; *p*=0.022) were associated with decreased risk of ILD endpoints. Among these proteins, only B3GALT6 showed strong evidence of genetic colocalization with hypothyroidism, as indicated by a posterior probability of colocalization exceeding 80%, with both associations driven by the same genetic variant (rs67492154) (**Figure 6B**;

**Supplementary Table S16**). No comparable colocalization evidence was observed for other proteins in relation to the respective diseases.

**Figure 6.**
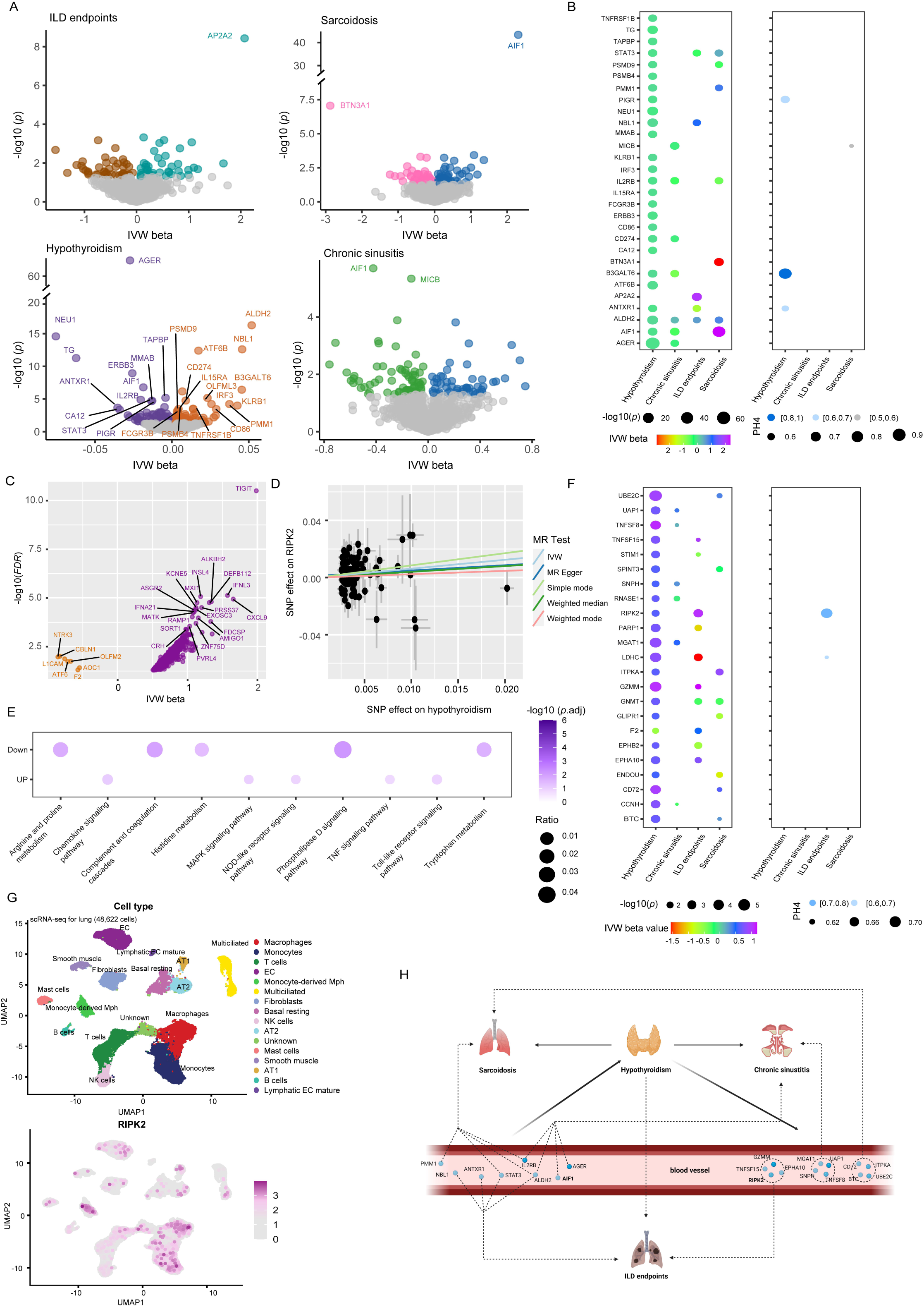
Prioritized plasma proteins associated with four diseases. (**A**) Volcano graphic depicting the effect of plasma protein on four diseases based on MR analyses. Color dots indicate significant relationships (uncorrected *p*<0.05). The labeled dot represents a significant association that passes multiple test corrections (FDR< 0.05). Only proteins with no significant heterogeneity (I^2^ < 50%), directional pleiotropy (*p-*_Egger intercept_ > 0.05), reverse causation, and passing leave-one-out analysis were presented in the plotting. (**B**) Shared the associations between 29 causal plasma proteins (statistically significant for at least one disease) and 4 diseases based on MR analysis. (Left) MR analysis reveals the causative effect of plasma proteins on four diseases; (right) colocalization analyses of *cis*-pQTL with four diseases. (**C**) Volcano plot depicting the effect of hypothyroidism on plasma protein based on MR analyses. Color dots indicate significant relationships after multiple testing corrections (FDR<0.05). Proteins were plotted with no significant heterogeneity (I^2^ < 50%), directional pleiotropy (*p-*_Egger intercept_ > 0.05), reverse causation, leave-one-out analysis, and multiple testing adjustments (FDR<0.05). (**D**) An MR scatter-plot depicting the effect of hypothyroidism on plasma RIPK2 levels. Using the inverse variance weighted method, an increase in hypothyroidism predicted by one standard deviation was linked to higher levels of RIPK2 (β=0.62, 95%CI=1.20-2.89, *p*=0.0052). (**E**) Enrichment analysis of biological functions for hypothyroidism-influenced proteins, both positively and negatively. (**F**) Shared the associations between 23 hypothyroidism-driven plasma proteins and diseases identified by MR analysis. (Left) Use of MR analysis to determine the causative influence of hypothyroidism-driven plasma proteins on diseases; (right) Colocalization analyses of *cis*-pQTL and four diseases. (**G**) Single-cell resolution analysis of RIPK2 expression in lung cell types from six healthy individuals and five systemic sclerosis-associated interstitial lung disease (SSC-ILD) patients. (Up) Sixteen annotated lung cell types. (Down) RIPK2 expression patterns in six healthy individuals and five SSC-ILD patients. UMAP, Uniform Manifold Approximation and Projection. EC, endothelial cell; AT, Alveolar Type. (**H**) A schematic illustration of the causes and consequences of proteins associated with hypothyroidism, as well as their relationship to the three diseases investigated. Figures generated with BioRender (https://biorender.com/).

### Identifying the causative effect of hypothyroidism-driven proteins on immune-related diseases

In fact, an alternative hypothesis is that circulating plasma proteins may mediate the effect of hypothyroidism on the development of immune-related diseases, thereby offering further insight into shared pathophysiological mechanisms^20^. To investigate this, a two-sample MR analysis was performed to estimate the causal influence of hypothyroidism on the abundance of 4,907 plasma proteins, treating hypothyroidism as the exposure and protein levels as the outcomes (**Figure 6C**; **Supplementary Table S17**). The F-statistic for the genetic instruments was 73.37, indicating robust instrument strength and minimal risk of weak instrument bias. After applying quality control measures, including exclusion based on heterogeneity (I^2^<0.5) and directional pleiotropy (MR-Egger intercept *p*>0.05), 1,315 proteins remained significantly associated with hypothyroidism (FDR<0.05). Proteins that failed the leave-one-out sensitivity analysis (n=207) or exhibited evidence of bidirectional causality (n=6) were subsequently excluded. This resulted in a final set of 1,102 hypothyroidism-driven proteins, including RIPK2, that met all robustness criteria: low heterogeneity (I^2^ < 50%), no directional pleiotropy, and absence of reverse causation (**Figure 6D**; **Supplementary Table S18**). The consistency of causal estimates for RIPK2 across multiple MR methods, including weighted median, weighted mode, and MR-Egger slope, further validated the IVW-based inference (**Supplementary Table S18**). Functional enrichment analysis of proteins positively regulated by hypothyroidism revealed significant overrepresentation in immune-related pathways, including Toll-like receptor signaling, chemokine signaling, NOD-like receptor signaling, and MAPK pathways. Conversely, proteins negatively affected were enriched in metabolic and neuroregulatory pathways, such as histidine and tryptophan metabolism, and the neurotrophin signaling pathway, highlighting the broad immunomodulatory and metabolic influence of hypothyroidism (**Figure 6E**).

To explore whether these hypothyroidism-regulated proteins contribute to the etiology of the associated immune-related diseases, a second two-sample MR analysis was subsequently conducted using *cis*-pQTLs as instrumental variables and GWAS data for ILD endpoints, sarcoidosis, and chronic sinusitis as outcomes. After harmonizing genetic instruments and outcomes, 130 proteins were evaluated. Although no associations survived FDR correction (FDR<0.05), 24 protein-disease pairs were nominally significant (**Figure 6F**). Among these, 13 showed effect directions consistent with mediation by hypothyroidism. For instance, elevated genetically predicted levels of RIPK2 were significantly associated with increased risk for ILD endpoints (OR=2.88 [95% CI: 1.55-5.38]; *p*=8.5×10^−4^). To assess potential confounding due to horizontal pleiotropy, RIPK2’s lead *cis*-pQTL (rs160438) was queried in the Open Targets Genetics platform (https://genetics.opentargets.org/). The variant showed no genome-wide significant associations with other traits in the deCODE dataset (*p*<5×10^−8^), supporting the specificity of the association. Colocalization analysis suggested moderate evidence for shared causal variants between RIPK2 and ILD endpoints (**Figure 6F**; **Supplementary Table S16**). To further investigate the cellular source of RIPK2 in lung tissue, single-cell RNA sequencing data were analyzed from samples obtained from six healthy individuals and five patients with systemic sclerosis-associated interstitial lung disease (SSC-ILD), as previously described^32,33^. RIPK2 expression was predominantly localized to monocytes, macrophages, and monocyte-derived macrophages (**Figure 6G**), indicating a potential role in myeloid-mediated immune responses within the lung microenvironment. Collectively, these findings support RIPK2 as a plausible mediator linking hypothyroidism to increased susceptibility to ILD endpoints.

## Discussion

In this study, we provide evidence for a shared genetic etiology between hypothyroidism and three immune-related diseases (namely sarcoidosis, chronic sinusitis, and ILD endpoints). These findings contribute to a deeper understanding of the genetic basis of comorbidity and may have implications for the elucidation of disease mechanisms and the identification of therapeutic targets.

Both GNOVA and LDSC analyses revealed significant genome-wide genetic correlations between hypothyroidism and the three immune-related diseases we studied, supporting the hypothesis that common genetic variants underlie their co-occurrence. By contrast, local genetic correlation analyses did not identify any specific genomic regions jointly contributing to the diseases, suggesting that the observed associations likely stem from polygenic overlap dispersed across the genome rather than localized hotspots. A significant causal relationship was found between hypothyroidism and two diseases (sarcoidosis and chronic sinusitis). Although a statistically significant association between hypothyroidism and ILD endpoints was also observed via IVW, this result was not replicated using the CAUSE model, indicating that the observed relationship may be confounded by correlated horizontal pleiotropy and should therefore be interpreted with caution.

Cross-trait meta-analyses and colocalization studies identified several shared genetic loci, strengthening the evidence for pleiotropy. Utilizing two complementary methods, namely MTAG and CPASSOC, we mitigated potential bias due to sample overlap and confirmed several consistently significant loci across both approaches. Notably, two novel shared variants were discovered: rs12806363, located in RPS6KA4, a kinase involved in serine/threonine signaling; and rs174573, a missense variant in FADS2, which encodes a fatty acid desaturase implicated in anti-inflammatory lipid metabolism. The increased expression of FADS2 in CD4⁺ T cells has been reported in asthma patients, highlighting its potential role involved in immune regulation^34^. Additionally, long-chain polyunsaturated fatty acids produced by FADS2 have anti-inflammatory effects and may regulate immunological function^35,36^. Further experiments are needed to investigate the role of FADS2 in the risk of hypothyroidism and chronic sinusitis disease.

Heritability enrichment analyses further revealed that central memory CD4⁺ T cells in these tissues, with strong colocalization evidence (posterior probability>0.9) supporting their involvement in the shared pathogenesis. This is consistent with recent report showing elevated levels of tissue-resident and effector memory CD4⁺ T cells in nasal polyps of patients with severe chronic rhinosinusitis^37^, suggesting a potential immunological link across these conditions. However, it is important to emphasize that heritability enrichment does not imply causality; future studies incorporating GWAS of tissue-resident immune cells are necessary to refine these findings.

To investigate potential molecular mediators, we integrated eQTL and pQTL datasets. Through SMR, HEIDI, and colocalization analyses, we identified CD226 and DOCK6 as putative genes linking hypothyroidism and sarcoidosis. CD226, a member of the immunoglobulin superfamily, is a functional protein originally produced on natural killer and T cells^38^. A recent study has reported that CD226 expression in T cells from sarcoidosis^39^. Furthermore, it was proposed that CD4+ T cells moving to the lungs via CXCR3 could be activated by CD226, potentially contributing to the pathogenesis of sarcoidosis. Similarly, DOCK6, a member of the DOCK-C family with GEF activity for Rac1 and CDC42, was identified as a shared gene based on data from both GTEx and eQTLGen. Recent research indicates that DOCK6 is linked to various diseases, including cancers like gastric cancer and acute myeloid leukemia, and functions as an oncogene^40–43^. However, its functional role in autoimmune or inflammatory conditions remains unclear and warrants further investigation. In parallel, we conducted a systematic evaluation of the proteomic landscape influenced by hypothyroidism. While no hypothyroidism-driven proteins showed significant causal effects on the three investigated diseases after correction for multiple testing, nominal associations were still observed in 24 protein-disease pairs. Notably, RIPK2 exhibited a suggestive mediating role in the relationship between hypothyroidism and ILD endpoints, supported by moderate colocalization evidence. Additional validation using *cis*-pQTLs in independent cohorts is still necessary to substantiate this finding.

Despite the strengths of this study, several limitations should be acknowledged. First, all GWAS datasets were derived from individuals of European ancestry, potentially limiting the generalizability of our findings to other populations. Second, the use of summary-level rather than individual-level data precluded stratified analyses by sex and age. Lastly, although we utilized high-quality GWAS datasets, more comprehensive datasets such as those used in the meta-analysis by Mathieu et al. (51,194 cases and 443,383 controls)^44^ may enhance discovery power. Nonetheless, our reliance on UK Biobank-based GWAS reduces potential bias from sample heterogeneity.

In conclusion, by integrating multi-omics data, our study identified significant genetic associations as well as the shared molecular components that link hypothyroidism to the other three immune-related diseases we studied. The identification of shared loci, genes, proteins, and immune cell types sheds new light on the genetic basis of disease comorbidities and identifies prospective targets for future therapeutic approaches. Additional functional validation is required to uncover the underlying mechanisms of these associations.

## Data Availability

All the datasets analyzed during the current study are publicly available.

## Acknowledgments

We thank Consortium des Equipements de Calcul Intensif en Fédération Wallonie Bruxelles (CECI) funded by the Fonds de la Recherche Scientifique de Belgique (FRS-FNRS) for providing the supercomputing facilities utilized for the analysis. The authors also thank all of the researchers involved in the associated genetic consortia and GWAS for making the data available to the public.

## Contributions

SF conceived and designed the study. SF and MJ performed the data analysis. SF and MJ conducted the interpretation of analysis results. SF wrote the manuscript. All authors read and approved the manuscript.

## Data and code availability

All the datasets analyzed during the current study are publicly available. Source data are provided with this manuscript.

## Material and methods

### GWAS data source

Summary-level data for the associations of hypothyroidism-associated SNPs were derived from the MRC-IEU Consortium (access id: ukb-b-4226), which involved 9,674 cases and 453,336 controls^16^. Summary-level data for interstitial lung disease endpoints (4,572 cases and 407,609 controls), sarcoidosis (4,399 cases and 405,620), chronic sinusitis (17,987 cases and 308,457 controls) were obtained from FinnGen study (R10 release)^17^. Summary statistics for 4,907 plasma proteins in 35,559 Icelanders were obtained from the Collaborative Analysis of Diagnostic Criteria in Europe project (deCODE)^30^. Summary data for 14 CD4+ memory-related T cells were retrieved from a sample of 3,757 Sardinians^28^. All *cis*-eQTL used in the present study were collected from both GTEx and the eQTLGen Consortium^26,29^. The UCSC tool liftOver was used to coordinate the genomic position of SNPs in the GWAS.

### Heritability and overall genetic correlation analysis

LD scores for approximately 1.2 million common SNPs were obtained from the HapMap3 reference panel, as calculated by the 1000 Genomes Project. Single-trait SNP-based heritability for hypothyroidism and three immune-related diseases, including ILD endpoints, sarcoidosis, and chronic sinusitis, was estimated using stratified linkage disequilibrium score regression (S-LDSC) under the baseline-LD model framework^45,46^. Heritability estimates were subsequently transformed to the liability scale based on both the observed sample prevalence and assumed population prevalence for each condition: 0.046 for hypothyroidism, 0.0003 for ILD endpoints, 0.000145 for sarcoidosis, and 0.08 for chronic sinusitis^47–50^. Pairwise genetic correlations were assessed using unconstrained LDSC, allowing the intercept to vary freely to account for potential sample overlap and population stratification^51^. Sensitivity analyses were also performed by constraining the LDSC intercepts, setting single-trait heritability intercepts to 1 and cross-trait intercepts to 0, based on the assumption of no sample overlap between the MRC-IEU Consortium and FinnGen datasets. A *p*-value threshold of 0.05 was applied to determine statistical significance. In parallel, genetic correlations were also evaluated using GNOVA, a genetic covariance analysis method known to provide more accurate and statistically powerful estimates of genetic correlation compared to LDSC^18^. GNOVA estimates genetic covariance by leveraging all overlapping SNPs across two GWAS summary statistics datasets and incorporates statistical correction for potential sample overlap between independent GWAS datasets. Analyses were conducted using the European reference panel from the 1000 Genomes Project, with default parameters applied.

### Local genetic correlation analysis

To assess the contribution of specific genomic regions to disease susceptibility, pairwise local genetic correlations were estimated using *ρ*-HESS with default parameters^18,23^. This method partitions the genome into 1,703 predefined LD-independent regions of approximately 1.5 Mb each, enabling region-specific evaluation of genetic associations. Statistical significance was defined using a Bonferroni-corrected threshold of *p*<0.05/1,703, while regions meeting a nominal significance level of *p*<0.05 were considered suggestive.

### Cross-trait GWAS meta-analysis

To investigate potential shared genetic risk factors between hypothyroidism and three immune-related diseases, cross-trait GWAS meta-analyses were performed to identify associated SNPs. Two complementary analytical approaches were employed: MTAG and CPASSOC methods^22,23^. MTAG increases statistical power by leveraging genetic correlations across multiple traits, offering an advantage over traditional single-trait GWAS. To evaluate the robustness of MTAG results, the upper bound of the false discovery rate (maxFDR) was calculated, providing an estimate of model validity under the assumption of equal variance-covariance structures for shared SNP effect sizes^15^. As a sensitivity analysis, CPASSOC was used to identify SNPs associated with at least one phenotype while accounting for potential confounding due to population structure or cryptic relatedness. CPASSOC employs the Shet statistic, which accommodates heterogeneous effect sizes across traits, thereby enabling more flexible modeling compared to MTAG. SNPs that reached genome-wide significance (*p*<5×10^−8^) in both MTAG and CPASSOC analyses (e.g., for hypothyroidism-sarcoidosis) but were not identified in the corresponding single-trait GWASs were prioritized. Independent loci were defined using LD clumping in PLINK (parameters: --clump-p1 5e-8, --clump-p2 1e-5, --clump-r2 0.2, --clump-kb 1000)^52^. Variants were considered novel if they were not in linkage disequilibrium (LD r^2^<0.2) with known lead SNPs from single-trait GWASs and were not primarily driven by a single phenotype. Finally, functional annotation of the identified variants was conducted using the Ensembl Variant Effect Predictor (VEP) to provide insights into their potential biological roles^53^.

### Summary data-based Mendelian randomization analysis

An analysis was performed using version 1.03 of the Summary-data-based MR (SMR) software^53^ to investigate the association between gene expression and four disease traits. Summary statistics from GWAS of hypothyroidism, sarcoidosis, chronic sinusitis, and ILD endpoints were integrated with eQTL data from GTEx (blood, lung, spleen, and small intestine) and eQTLGen (blood). Significant associations between gene expression and disease outcomes were determined using a Bonferroni-corrected significance threshold, defined as SMR *p*<0.05 divided by the number of genes tested per trait. To assess whether the observed associations were due to pleiotropy or linkage, HEIDI tests were applied. A *p-*_HEIDI_<0.05 indicates that the association may be driven by distinct causal variants in linkage disequilibrium, rather than a shared causal variant affecting both gene expression and disease risk.

### Colocalization analysis and Bayesian fine-mapping analysis

Colocalization analysis was conducted to assess whether hypothyroidism and the associated complex diseases share causal genetic variants within specific genomic regions. This was implemented using the “coloc.abf” function from the coloc R package, following previously established methodologies^54,55^. The approach employs a Bayesian framework to estimate posterior probabilities for five mutually exclusive hypotheses regarding the presence and sharing of causal variants across traits. A posterior probability for hypothesis 4 (PH4) exceeding 0.8 was interpreted as strong evidence of colocalization, indicating a shared causal variant^54^. In parallel, fine-mapping of associated loci was carried out using the “susie_rss” function from the susieR R package (version 0.11.42)^24^. This method generated 95% credible sets of SNPs likely to contain the true causal variant for each independently associated region, defined within a 500-kilobase (kb) window.

### Tissue enrichment analysis

To identify tissues most relevant to disease phenotypes, tissue-specific enrichment analyses were conducted using the GTEx v8 dataset in combination with LDSC and MAGMA, following the framework established by Bryois et al.^56^. Briefly, median gene expression values across individuals were utilized, with exclusions applied to non-biological tissues, testis, and any tissue type sampled in fewer than 100 individuals. Expression values were then aggregated by organ system (excluding brain tissues), resulting in expression profiles for 37 distinct tissues. For downstream analyses, the top 10% of genes exhibiting the highest tissue specificity in each tissue were selected. In the LDSC framework, SNPs located within 100-kb windows surrounding these highly tissue-specific genes were independently incorporated into the baseline-LD model for each tissue. The significance of enrichment was evaluated using the z-score of the regression coefficient. For MAGMA, gene-level association statistics were calculated using a genomic window extending 35 kb upstream and 10 kb downstream of each gene, based on the European population reference from phase 3 of the 1000 Genomes Project. MAGMA was used to assess whether the top 10% most tissue-specific genes in each tissue showed enrichment for genetic association with the traits of interest. For both LDSC and MAGMA analyses, statistical significance was determined using a FDR threshold of 5% across all tissues evaluated per trait.

### Cell-type enrichment analyses using scRNA-seq datasets

To identify cell types potentially contributing to the genetic architecture of complex traits, scRNA-sequencing data from four tissues, including whole blood^57^, spleen^58^, small intestinal^59^, and lung^60^, were integrated with GWAS summary statistics for hypothyroidism and three related diseases. This integration was performed using three complementary gene prioritization frameworks: linkage disequilibrium score regression applied to specifically expressed genes (LDSC-SEG), MAGMA, and scDRS^25,27,61^. LDSC-SEG and MAGMA analyses were conducted using the CELLECT Snakemake workflow^62^. Within this framework, cell-type-specific expression scores (ESm) were computed for each cell population using the CELLEX method. SNP annotations were generated using data from the 1000 Genomes Project, consistent with the baseline model for stratified LDSC (S-LDSC)^63^. LD scores were calculated using HapMap3 SNPs, excluding the major histocompatibility complex (MHC) region due to its complex LD structure. Subsequent enrichment analyses were performed using both LDSC-SEG and MAGMA regression approaches. Concurrently, scDRS was employed to link single-cell gene expression profiles with polygenic GWAS data, identifying cell subpopulations enriched for disease-associated genetic signals. Disease-relevant gene sets were constructed using the top 1,000 MAGMA-prioritized genes, ranked by Z-score. scDRS scores were computed via the CLI command “scdrs compute-score”, incorporating a covariate matrix that included assay type, sex, age, ethnicity, and, when available, the duration of cold storage. To control for multiple hypothesis testing, FDR correction was applied to all *p*-values across tissues and diseases using the Benjamini-Hochberg procedure

### Mendelian randomization and mediation analysis

Two-sample MR analyses were conducted to investigate potential causal relationships, employing the TwoSampleMR R package (version 0.5.6) and the CAUSE framework, as previously described^55^. Briefly, SNPs reaching genome-wide significance (*p*<5×10^−8^) and located outside the MHC region (chromosome 6: 28,477,797–33,448,354, GRCh37) were selected. Independent instrumental variables (IVs) were identified based on LD criteria (r^2^<0.001, clumping window more than 10,000 kb) using the 1000 Genomes Project European reference panel. The analysis focused on *cis*-pQTLs, defined as variants within 500 kilobases upstream or downstream of the gene body, to explore plasma proteins potentially associated with disease outcomes. When two or more valid IVs were available for a protein, the IVW method served as the primary analytical approach; for proteins with only one instrument, the Wald ratio was applied. To evaluate horizontal pleiotropy, the MR-Egger intercept test was performed, while heterogeneity among instruments was assessed using Cochran’s Q statistic and I^2^, calculated via the “Isq()” function. Leave-one-out sensitivity analyses were conducted to identify any single IV driving the observed association. To further mitigate potential pleiotropic bias, each SNP was cross-referenced with the Open Targets Genetics database (https://genetics.opentargets.org/) to identify previously reported associations. SNP-disease associations with *p*<5×10^−8^ were considered statistically significant. To adjust for multiple comparisons, the Benjamini-Hochberg procedure was applied, and associations were considered significant at a FDR threshold of <0.05. Additionally, the CAUSE model was employed to account for both correlated and uncorrelated horizontal pleiotropic effects. CAUSE uses a multivariate framework that models the joint distribution of SNP-exposure and SNP-outcome associations to reduce false positives arising from correlated pleiotropy^21^. To quantify mediation effects (β_mediated_) network MR was implemented using the product-of-coefficients method, following the approach described by Yoshiji et al.^20^. Specifically, we estimated the effect of protein levels on hypothyroidism (β_protein-to-hypothyroidism_) and the effect of hypothyroidism on chronic sinusitis (β_hypothyroidism-to-chronic sinusitis_). The mediated effect was calculated as β_mediated_=β_protein-to-hypothyroidism_×β_hypothyroidism-to-chronic sinusitis_) and the proportion mediated was determined by dividing β_mediated_ by the total effect β_protein-to-chronic sinusitis_.

## Supplementary Figure

**Figure S1. Local genetic correlations between hypothyroidism and three complex diseases revealed by ρ-HESS (Heritability Estimation from Summary Statistics).** Local genetic correlations between hypothyroidism and chronic sinusitis (A), ILD endpoints (B), and sarcoidosis (C), respectively. Significant local genetic correlation estimates are highlighted in red and blue for even and odd chromosomes, respectively.

## Supplementary Table

**Supplementary Table S1**. Genetic correlations between hypothyroidism and complex diseases.

**Supplementary Table S2**. Summary of localized genetic correlations between hypothyroidism and complex diseases.

**Supplementary Table S3**. Summary of Mendelian randomization results between hypothyroidism and complex diseases using GWASs of hypothyroidism from MRC-IEU Consortium.

**Supplementary Table S4**. Summary of Mendelian randomization replication results between hypothyroidism and complex diseases using GWASs meta-analysis of hypothyroidism.

**Supplementary Table S5**. Genome-wide significant loci in a genome-wide cross-trait analysis approach.

**Supplementary Table S6**. Shared risk regions in different analyses.

**Supplementary Table S7**. List of 95% credible set SNPs for each independent locus using fine mapping analysis.

**Supplementary Table S8**. Genetic enrichment for hypothyroidism and complex diseases in specific tissues using LDSC and MAGMA.

**Supplementary Table S9**. Genetic enrichment for hypothyroidism and complex diseases in cell-type using LDSC, MAGMA, and scDRS.

**Supplementary Table S10**. Summary of Mendelian randomization results for causal effects of CD4+ memory-related cells on diseases.

**Supplementary Table S11**. Summary of Mendelian randomization results for the causal effect of diseases on CD4+ memory-related cells.

**Supplementary Table S12**. SMR-prioritized genes associated with hypothyroidism and three complex diseases.

**Supplementary Table S13**. Co-localization analysis of shared SMR priority genes in Table S12.

**Supplementary Table S14**. Analyzing the causal effect of plasma proteins on disease using MR (unfiltered).

**Supplementary Table S15**. Significant causal effects of plasma proteins on disease.

**Supplementary Table S16**. Co-localization analysis of etiological proteins that have a causal impact on disease.

**Supplementary Table S17**. Causal effect of hypothyroidism on plasma proteins using MR (unfiltered).

**Supplementary Table S18**. Significant causal effect of hypothyroidism on plasma proteins (hypothyroidism-driven proteins).

